# Genomic epidemiology of SARS-CoV-2 in Russia reveals recurring cross-border transmission throughout 2020

**DOI:** 10.1101/2021.03.31.21254115

**Authors:** Alina Matsvay, Galya V. Klink, Ksenia R. Safina, Elena Nabieva, Sofya K. Garushyants, Dmitry Biba, Georgii A Bazykin, Ivan M. Mikhaylov, Anna V. Say, Anastasiya I. Zakamornaya, Anastasiya O. Khakhina, Tatiana S. Lisitsa, Andrey A. Ayginin, Ivan S. Abramov, Sergey A.Bogdan, Kseniya B. Kolbutova, Daria U Oleynikova, Tatiana F. Avdeenko, German A. Shipulin, Sergey M.Yudin, Veronika I. Skvortsova

## Abstract

SARS-CoV-2 has spread rapidly across the globe, with most nations failing to prevent or substantially delay its introduction. While many countries have imposed some limitations on trans-border passenger traffic, the effect of these measures on the spread of COVID-19 strains remains unclear. Here, we report an analysis of whole-genome sequencing of 3206 SARS-CoV-2 samples from 78 regions of Russia covering the period between March and November 2020. We describe recurring imports of multiple COVID-19 strains throughout this period, giving rise to 457 uniquely Russian transmission lineages, as well as repeated cross-border transmissions of local circulating variants out of Russia.

---

The earliest known case of COVID-19 was admitted to a hospital in Wuhan, China, on December 16, 2019^1^, and the first case outside China was reported on January 13, 2020^2^. In Russia, the first case was reported on March 1, 2020 in a patient returning from Italy^3^. By the time Russia closed its borders on March 30, 2020, 1836 cases were reported in Russia^4^ and 888,460 globally^5^. SARS-CoV-2 has spread rapidly throughout Russia, with evidence for tens of introductions by summer 2020^6,7^. COVID-19 cases in Russia have spiked in May and then again near the end of 2020, with a total of 3,612,800 documented cases as of January 19, 2021^5^.

After border closure in March 2020, international air traffic from and to Russia was reduced from four-five million passengers per month in January-February to ∼30,000 in April. The border was gradually reopened starting from early August^8^; accordingly, numbers of travellers increased, reaching ∼1.5 million per month in September and October 2020^9^.

Here, we detail the dynamics of COVID-19 variants in Russia between March-November 2020. We show that the intensity of cross-border transmission remained high throughout this period, with multiple transfers of variants in both directions.

## Diversity of SARS-CoV-2 in Russia

To study the diversity of SARS-CoV-2 in Russia, we sequenced complete viral genomes from 1620 samples obtained between March 02 and November 25, 2020. We combined this data with the 1570 genomes from Russia for the period of March 11 - November 28 that we downloaded from GISAID on January 5th, 2021. We also included one sample obtained in Japan on February 25 from a Diamond Princess passenger who later returned to Russia; and 15 samples from Kazakhstan obtained from Russian citizens. The resulting dataset included 3206 sequences from 78 out of the 85 regions of the Russian Federation (Fig. 1, 2).

**Fig. 1.**
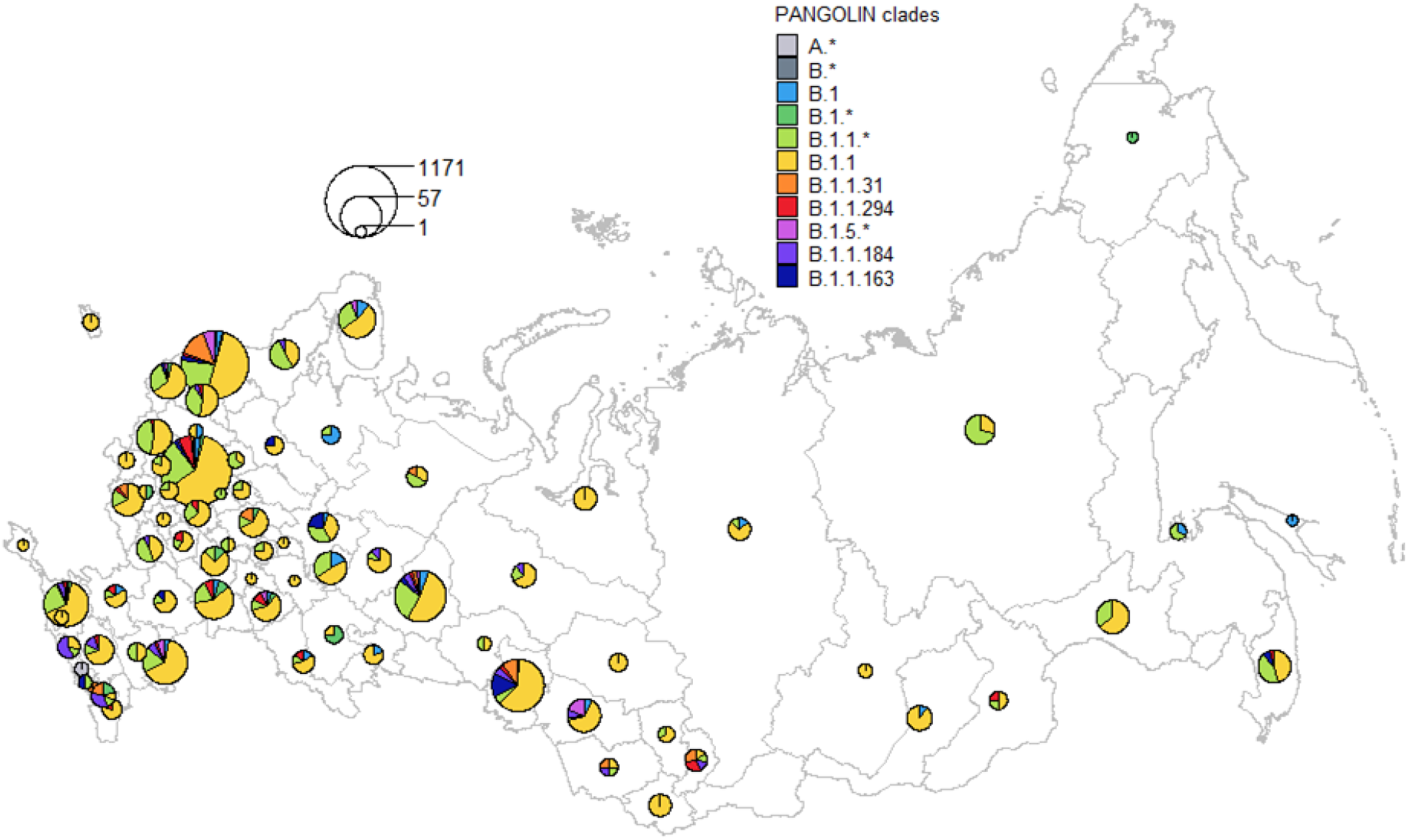
Prevalence of major PANGOLIN lineages^10^ in Russia. The data presented is for all sequences collected between March-November, 2020; see Fig. S1 for a breakdown by period. The circle size is proportional to the number of samples in corresponding regions, categorized by PANGOLIN lineages. Moscow is pooled with the surrounding Moscow Region, and Saint Petersburg is pooled with the surrounding Leningrad Region. Asterisks in PANGOLIN lineage designations correspond to pooled sets of lineages of that hierarchy level, except those listed in other categories; e.g., B.1.1.* includes B.1.1.7 but not B.1.1 or B.1.1.31.

**Fig. 2.**
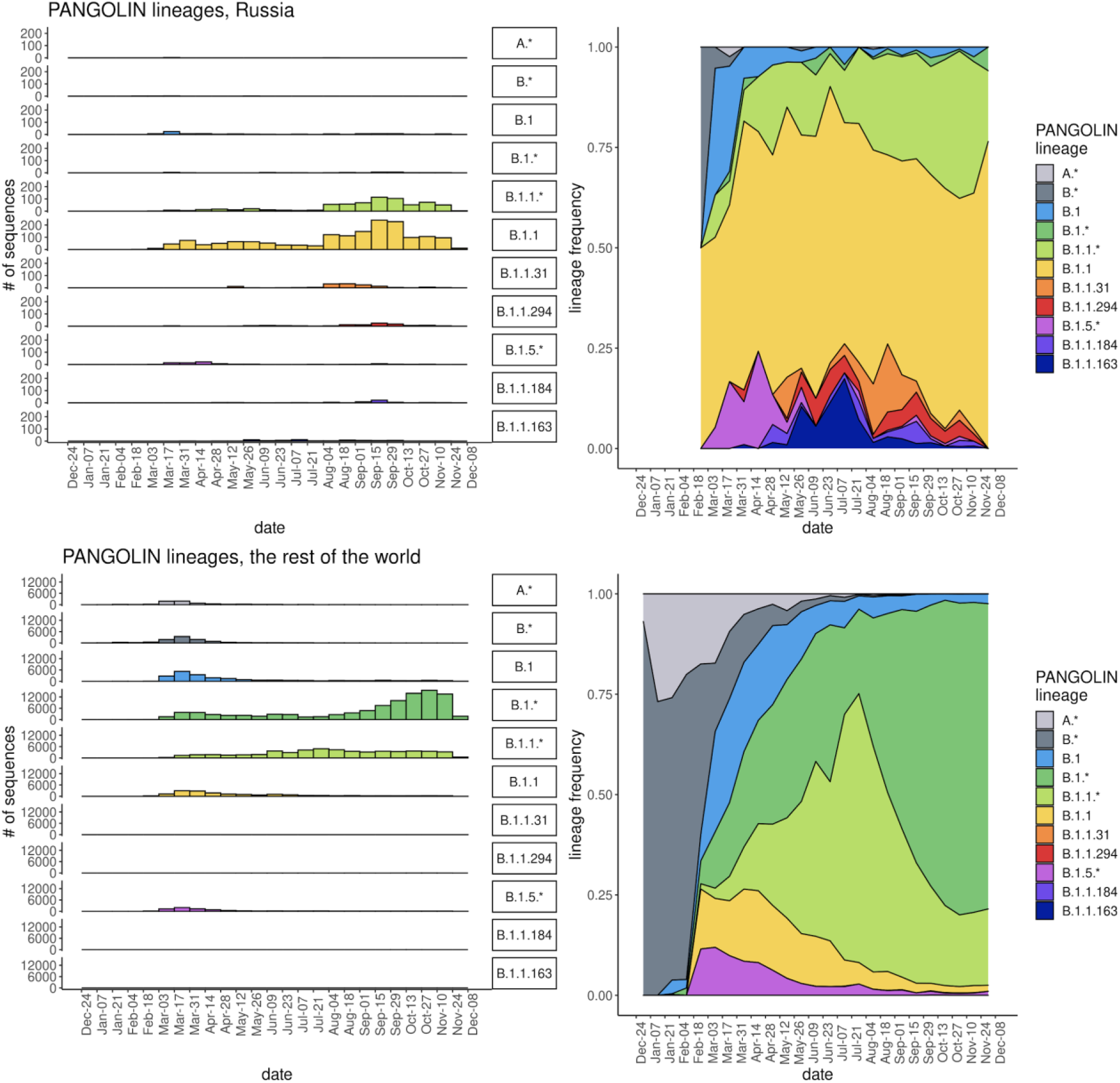
Dynamics of PANGOLIN lineage frequencies in Russia (top row) and in the rest of the world (bottom row). Sequences are ordered by sampling date, split into 14 day bins. Color legend as in Fig. 1.

To study the origin of SARS-CoV-2 in Russia in more detail, we reconstructed a maximum likelihood phylogeny of SARS-CoV-2 that comprised 3,206 Russian and 217,319 non-Russian sequences. Following previous work^6,11^, we then grouped Russian samples into Russian transmission lineages on the basis of their phylogenetic positions. Each Russian transmission lineage carries one or more characteristic changes at its base. A single introduction of a viral strain into Russia can result in one or more Russian transmission lineages (Fig. S2a). The remaining samples were as much or more related to non-Russian sequences as to the Russian ones, and were therefore not grouped into Russian transmission lineages. Such samples were classified as “singletons” if they carried characteristic changes of their own, or as “stem clusters” if they did not^6^ (Fig. S2a-b).

We identified 457 uniquely Russian transmission lineages, together encompassing 2089 (65%) of the sequenced samples. The remaining sequences represented singletons (11%) or stem clusters (24%). Many of the PANGOLIN lineages, notably the B.1.1 lineage, were brought into Russia repeatedly (Fig. 3). The earliest sampling dates for Russian transmission lineages fall throughout the study period (Fig. 3). The number of Russian transmission lineages continues to increase with the number of obtained sequences with little evidence for saturation (Fig. S3), suggesting that some lineages remain undetected and/or that lineages continue to be imported.

**Fig. 3.**
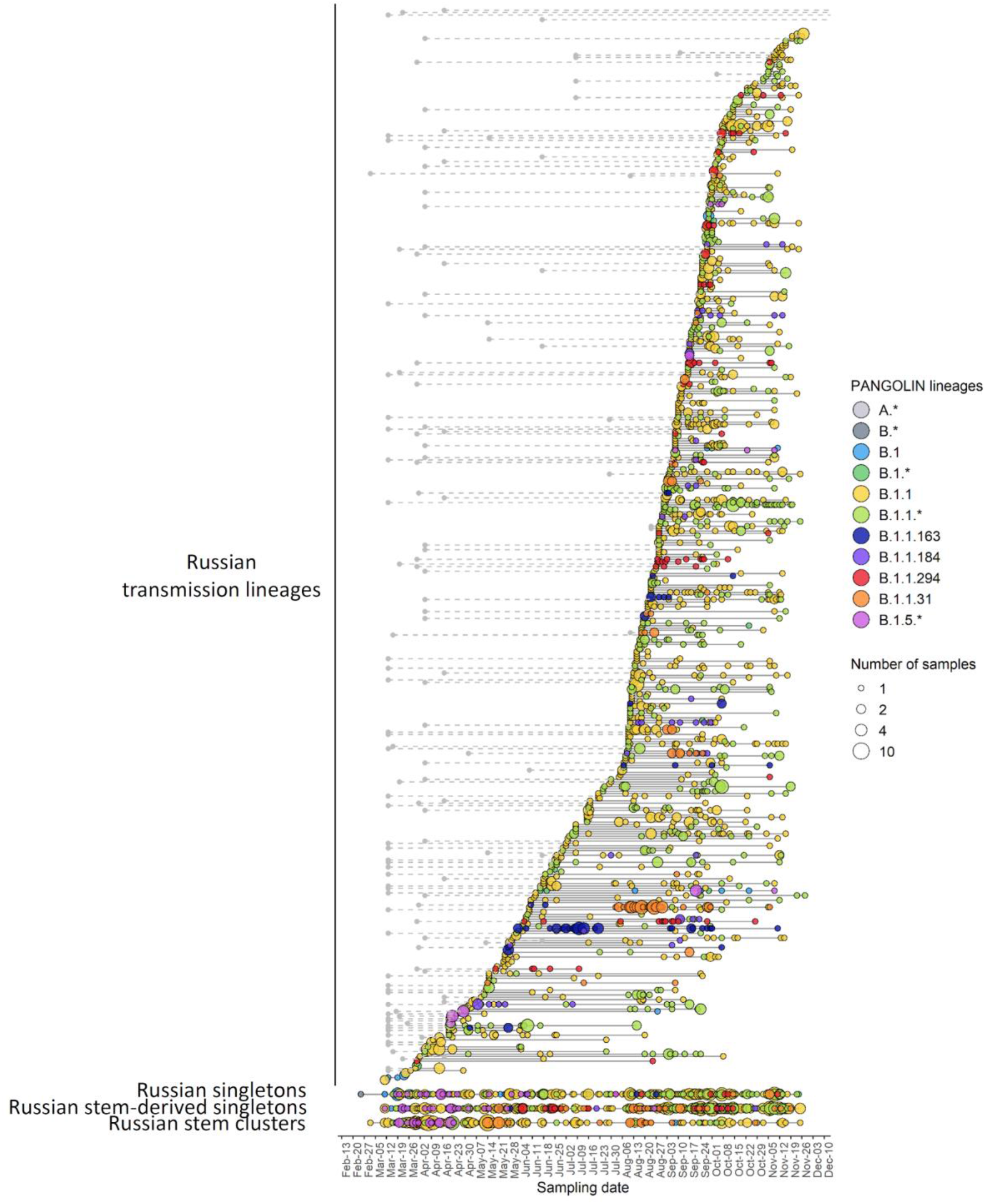
Russian transmission lineages. In the top part of the figure, each horizontal line represents a Russian transmission lineage, ordered by the date of the earliest sample. For Russian transmission lineages descendant from a Russian stem cluster, the grey circle identifies the earliest sequence date for the corresponding stem cluster. The three bottom lines represent Russian singletons, Russian stem-derived singletons, and Russian stem clusters (see Fig. S2 for terminology for lineage notation). Circles represent samples taken on a particular date, with circle size representing the number of samples. Circle color indicates the PANGOLIN designation of the corresponding sample.

Overall, Russian transmission lineages were rather well mixed within Russia. 216 of the 454 lineages with known sample locations (47,6%) were each observed in two or more regions, suggesting intensive transport of individual lineages within Russia (Fig. 4). The remaining 238 lineages were limited in their spread to just one region.

**Fig. 4.**
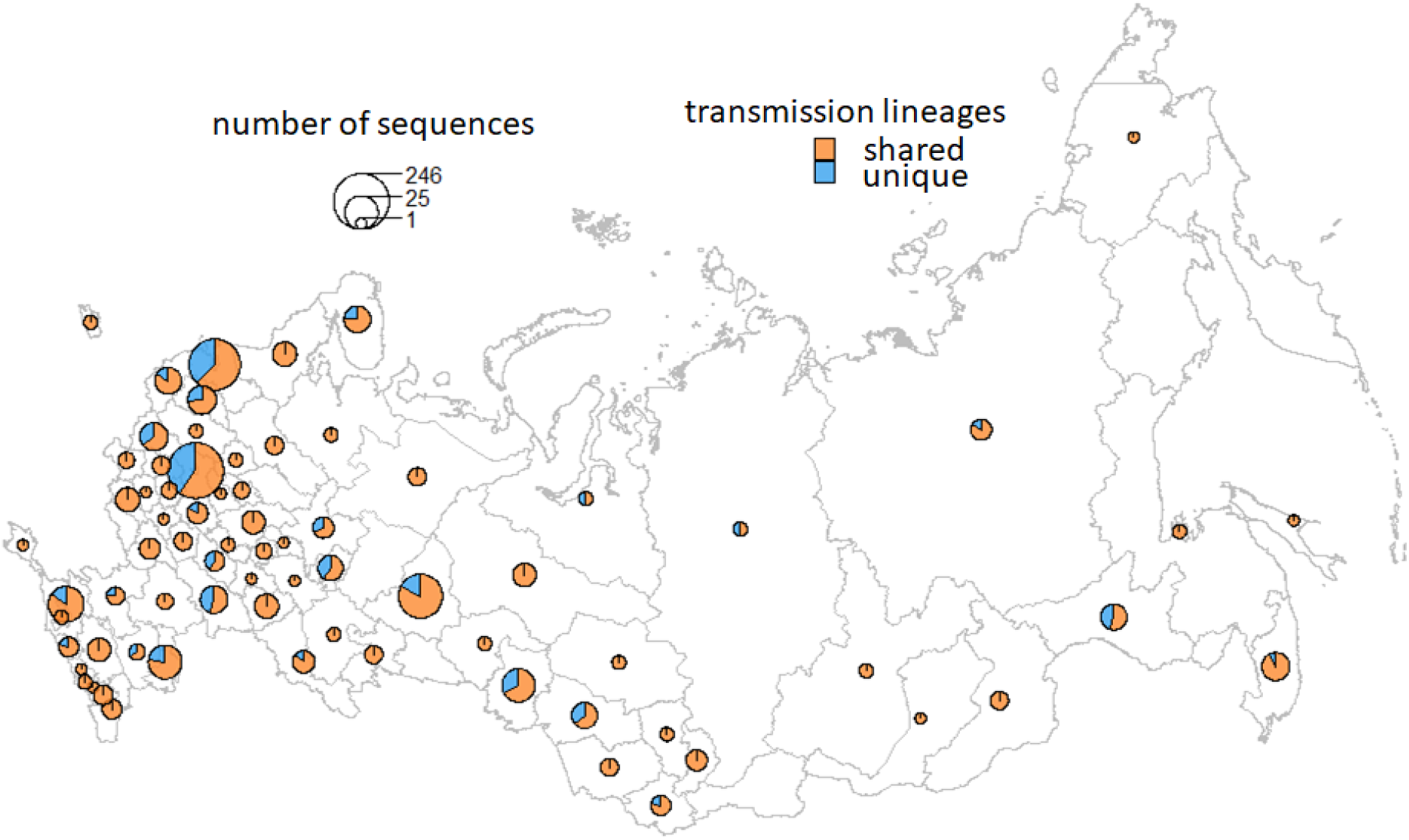
Fractions of Russian transmission lineages unique to one region or shared with other regions. Circle size is proportional to the number of Russian transmission lineages in the corresponding region.

## Cross-border transmission of SARS-CoV-2 strains

We asked how SARS-CoV-2 lineages were imported into Russia (inbound transmissions, IBT) and transmitted out of it (outbound transmissions, OBT). To obtain conservative estimates for numbers of IBTs and OBTs, we introduced stringent definitions for these events, defining all sister Russian lineages descendant from a non-Russian ancestor as results of a single IBT, and all sister non-Russian lineages descendant from a Russian ancestor, as results of a single OBT (see Methods, Fig. S2c-d). Under these definitions, we observe a total of 82 IBTs, and 43 OBTs. These numbers are the lower boundaries; in particular, multiple Russian transmission lineages derived from a single stem node are here considered a result of a single IBT. An alternative definition of IBTs and OBTs obtained using Treetime^12^ (Figure S4; see Methods) yielded larger numbers of inferred IBTs (418) and OBTs (118); nevertheless, these lists included 94% (118 of 125) of events detected using our definition, and all of the seven events absent in the Treetime-based list could be also inferred from the Treetime output with a lower probability threshold (0.75 instead of 0.8; see Methods), indicating robustness of inferred IBTs and OBTs.

Overall, we find that transmissions of SARS-CoV-2 variants into Russia which started in spring continued throughout summer and fall, in spite of interventions aimed at curbing transborder travel. Similarly, we observe likely transmissions out of Russia starting in spring, and continuing throughout the summer and fall (Fig. 5).

**Fig. 5.**
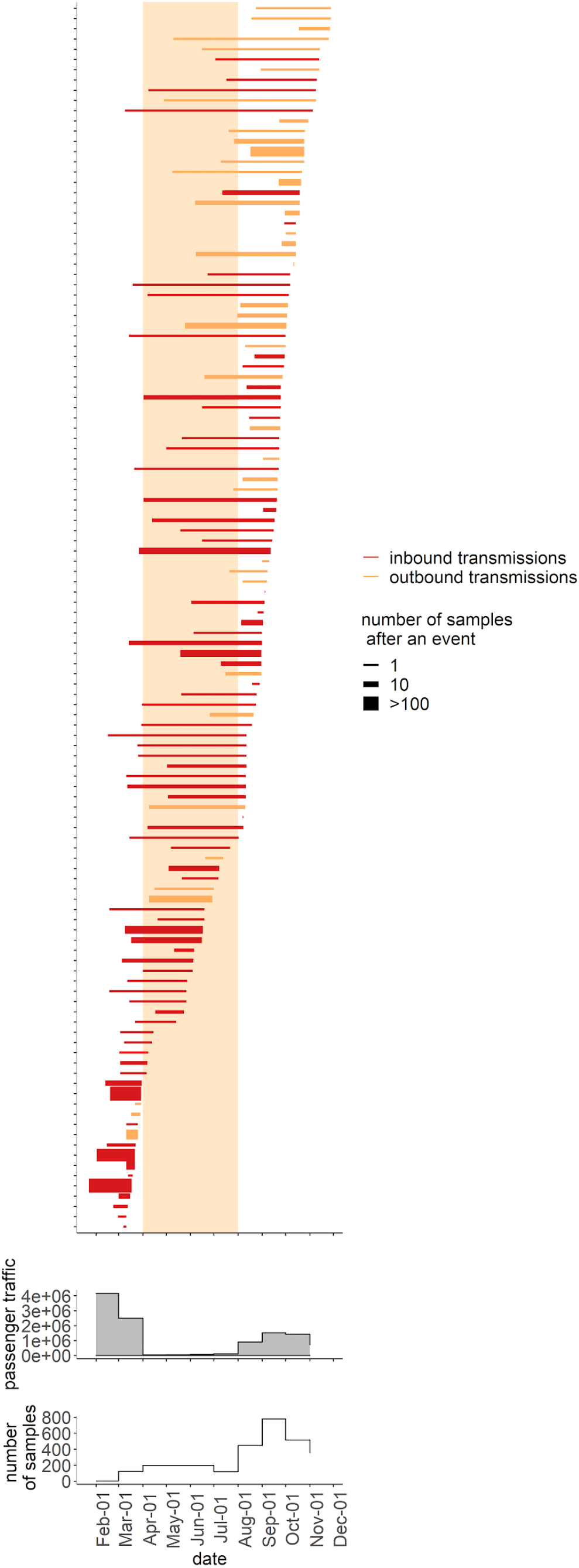
Timeline for inbound and outbound transmissions of SARS-CoV-2 variants. Bars correspond to individual IBTs (red) and OBTs (orange), with the left end of the bar corresponding to the date estimated for the corresponding node by the “node.dating” procedure^13^ from the “ape” package of R language^27^, and the right end of the bar corresponding to the date of the earliest Russian (for IBTs) or non-Russian (for OBTs) sample among the node’s descendants. Line width identifies the number of sequences descendant to the IBT or OBT. Only cases with available sample dates are shown. Also shown is the number of international travellers going through Russian airports and the total number of Russian samples obtained in the corresponding month.

For a fraction of phylogenetically inferred cross-border transmissions, it was possible to obtain independent confirmation. These were OBTs to countries with low overall number of cases (allowing cross-referencing of phylogenetic and epidemiological data) that sequence intensively; for example, New Zealand sequences and shares openly ∼48% of its covid cases^14^. We were able to cross-reference five OBT events in this way, detailed below.

Two transmissions from Russia to New Zealand, giving rise to a total of 12 samples (Fig 6; Fig S5), are associated with mariners who travelled by a charter flight from Russia and Ukraine in the middle of October^15,16^. A total of thirty-three cases in New Zealand are linked to this group, including two staff members of the isolation facility^17^. We show here that these mariners have brought with them at least two distinct variants of SARS-CoV-2.

**Fig. 6.**
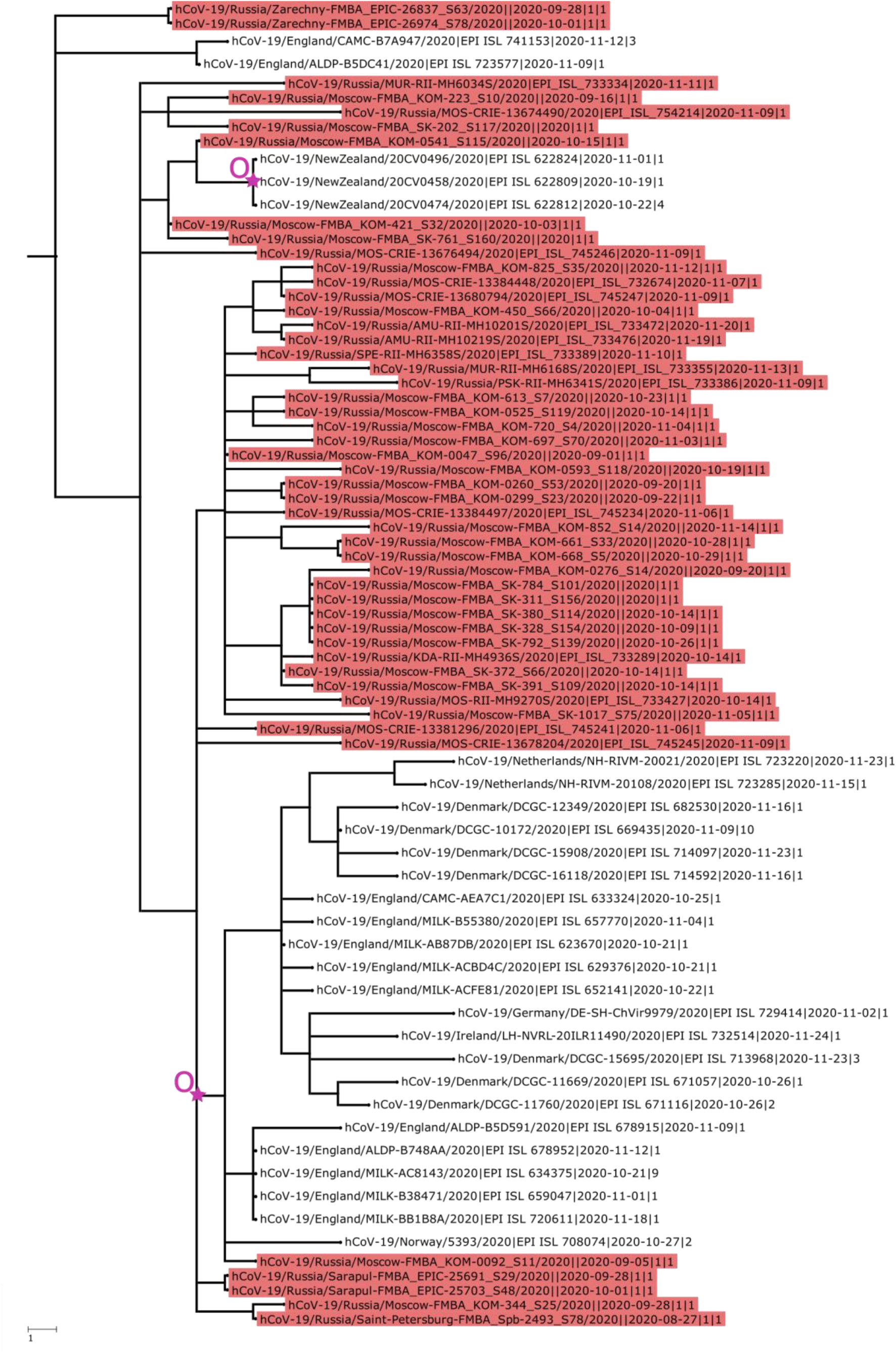
IBTs and OBTs in the history of the B.1.1.238 lineage. Branch lengths are measured in the number of changes. The number at the end of the sequence id represents the number of identical sequences (including the one shown) identified in the region on this date. Samples from Russia are identified with red labels. OBTs are indicated by a purple star and “O” letter.

Additionally, a late June sample from South Korea is nested within a Russian clade together with a number of UK samples (Fig. S6a), signifying an OBT. Unfortunately, no detailed information on the sources of the British sequences is available, but the sample from South Korea is marked as imported in the GISAID metadata. We were not able to specify this further, although according to media reports, multiple ships with Russian mariners who tested positive for SARS-CoV-2 were located at the time in the port of Busan^18^.

Finally, there are two more clusters of sequences from South Korea that group with Russian sequences (Fig. S6b,c). The first includes one sequence dated October 14, and the second, two sequences dated October 30. All these sequences are marked as imported from Russia in GISAID metadata.

Availability of genetic sequences from different countries allows to infer international transmission of COVID-19 lineages. Here, we nearly double the amount of whole-genome sequencing data for SARS-CoV-2 from Russia. Using genomic epidemiology, we show that genetically distinct SARS-CoV-2 variants have been imported into Russia at least 82 times, resulting in several hundreds of Russian transmission lineages. Similar to many other countries, we also detect outbound transmission of SARS-CoV-2 variants, and describe 43 putative outbound transmission events.

We observe intensive transmission of lineages both into Russia (IBTs) and outside it (OBTs). For a small fraction of samples, the direction of OBTs could be identified using public data. While these were the cases when the OBTs were stopped at the border, some of them resulted in onward transmission in corresponding countries, highlighting the public health relevance of these events.

While a detailed analysis of the effect of transit limitations on cross-border transport of lineages is beyond the scope of this study, our analysis suggests that IBTs and OBTs of lineages were not fully stopped by a radical reduction of the air passenger flow. Epidemiological simulations show that limiting the number of flights and/or cancelling direct flights between two countries may not be sufficient to prevent transmission of variants between them^19^. Even weak passenger flow from a country with high prevalence of COVID-19 and high frequency of a variant may be sufficient for its import^11^, and relatively few imports may suffice to cause sustainable domestic transmission, especially if these variants confer selective advantage for the virus, e.g. due to increased transmissivity^19^. The recent emergence of multiple transmissivity-increasing variants calls for a systematic and effective approach to curbing their global spread.

## Methods

### Sample collection

Nasopharyngeal swabs from patients with confirmed COVID-19 infection were collected between March 02 and November 25, 2020 by clinical hospitals subordinated to the Federal Medical-Biological Agency (FMBA) of Russia. All samples were collected to the transport media (Single-Use Virus Collection Tube, CDVCT-1, CDRICH) suitable for SARS-Cov-2 virus storage, transferred to the laboratory within 48 hours after collection and stored at −80°C prior to RNA extraction and sequencing. Transportation and storage were compliant with the local normative documents for handling of biologically hazardous samples.

For each sample, the following data were stored: unique identifier, date of sample collection, and geographic region of origin. The obtained samples covered the following regions of Russia: Altai Region, Amur Region, Arkhangelsk Region, Astrakhan Region, Chelyabinsk Region, Chukotka Region, Chuvash Region, Irkutsk Region, Ivanovo Region, Kaluga Region, Kamchatka Region, Khabarovsk region, Khanty-Mansi Region, Kirov Region, Krasnodar Region, Krasnoyarsk Region, Kursk Region, Leningrad Region, Moscow, Moscow Region, Murmansk Region, Nizhny Novgorod Region, Novosibirsk Region, Penza Region, Perm Region, Primorsky Region, Rostov Region, Ryazan Region, Saint Petersburg, Sakha Region, Samara Region, Saratov Region, Smolensk Region, Stavropol Region, Sverdlovsk Region, Tomsk Region, Tver Region, Tyumen Region, Udmurtia Region, Ulyanovsk Region, Volgograd Region, Voronezh Region, and Zabaykalsky Region.

### Whole-genome sequencing

The presence of SARS-CoV-2 RNA was confirmed using the *AmpliTest* SARS-CoV-2 test kit; samples with Ct (Hex) below 25 were considered positive. RNA from positive samples was isolated using the Ribo-prep purification kit and reverse-transcribed using the Ampliseq cDNA Synthesis for Illumina kit (Illumina; San Diego, CA, USA)). The resulting cDNA was amplified using the AmpliSeq for Illumina SARS-CoV-2 Research Panel (Illumina) which contains 247 amplicons in 2 pools targeting the whole SARS-CoV-2 genome. Library preparation was performed using the AmpliSeq Library PLUS kit (Illumina). Library quality was assessed by capillary electrophoresis using the Agilent 2100 Bioanalyzer system (Agilent; Santa Clara, CA, USA). Library concentration was measured with the Qubit 4 Fluorometer (Thermo Fisher Scientific; Waltham, MA, USA) using the Qubit dsDNA HS Assay Kit (Thermo Fisher Scientific). Sequencing was carried out on the Illumina NextSeq 550 System with the NextSeq 500/550 Mid Output Kit v2.5 (300 Cycles) (Illumina). The manufacturers’ recommendations were followed in all cases.

### Consensus calling

Paired-end sequencing data were generated for a total of 2,046 samples. After combining reads from the same samples, 1,903 unique sequenced samples remained. These sequences were then subjected to the following combination of read trimming and filtration: raw reads were trimmed with Trimmomatic-0.39 (ref. ^20^) to remove adapter sequences and low-quality ends. Trimmed reads were mapped onto the Wuhan-Hu-1 (MN908947.3) reference genome with bwa mem^21^. The following reads were then removed from the mapping: reads with abnormal insert length-to-read ratio (greater than 10 or less than 0.8); reads with insert length greater than 600; reads with more than 50% soft-clipped bases. 25 nucleotides were cropped from read ends using custom scripts to get rid of potential primer sequences. SNV and short indel calling was done with lofreq^22^, with SNVs considered consensus if they were covered by at least 4 reads and supported by more than 50% of those reads; indels were considered consensus if they were covered by at least 20 reads with at least 50% of those supporting the variant. Regions that were covered by fewer than 2 reads or that were covered by 2 or 3 reads and called non-reference were masked as N. Sequences with more than 3,000 nucleotides marked as N were removed from further analysis, resulting in 1,636 sequences. Consensus sequences were generated by bcftools consensus^23^.

### Data preparation and filtering

321,100 genomes of SARS-CoV-2 were downloaded from GISAID on January 6, 2021, (Supplementary Data ACKN) and aligned with MAFFT v7.453^24^ against the reference genome Wuhan-Hu-1/2019 (NCBI ID: MN908947.3) with --addfragments - -keeplength options. 100 nucleotides from the beginning and from the end of the alignment were trimmed. After that, we excluded sequences (1) shorter than 29,000 bp, (2) with more than 3,000 (for Russian sequences) or 300 (for all other countries) positions of missing data (Ns), (3) excluded by Nextstrain^25^, (4) from animals other than minks, (5) with a genetic distance to the reference genome more than four standard deviations from the epi-week mean genetic distance to the reference, or (6) obtained later than the most recent Russian sample (2020-11-28), leaving us with 218,889 sequences. To this dataset, we added the 1,636 sequences produced in this study. For all countries but Russia, closely-related sequences were then collapsed within the country using cd-hit with -c 0.99996 option. The final dataset comprised of 123,432 sequences was then annotated by the PANGOLIN package (v2.1.6, released on 2020-01-02) and split into five subsets corresponding to the major distinct SARS-CoV-2 clades: A, B.x, B.1.x, B.1.GH and B.1.1 (see Fig. S7). We additionally masked a highly homoplasic site 11,083 prior to reconstructing phylogenies.

### Phylogenetic analysis

For each of the five subsets, we constructed a phylogenetic tree using IQ-Tree v2.1.1^26^ under the GTR substitution model and ‘-fast -altr 1000’ as options and reconstructed ancestral sequences at the internal tree nodes with TreeTime v0.8.0^12^. All subsequent analyses were performed on each dataset independently. Samples were grouped into Russian transmission lineages as described in ref. ^6^, with the difference that we no longer distinguished between Russian transmission lineages and Russian stem-derived transmission lineages. Russian stem clusters, Russian singletons and Russian stem-derived singletons were inferred as described in ref. ^6^.

Inbound transmissions (IBTs) and outbound transmissions (OBTs) were defined as follows. A node I is assumed to be ancestral to an IBT event if (i) all descendants of the node ancestral to I (other than I) are non-Russian; (ii) one or more of immediate descendants of I are clades consisting of only non-Russian sequences, and (iii) one or more of immediate descendants of I are Russian singletons or Russian transmission lineages (Fig S2c). Analogously, a node O is assumed to be ancestral to an OBT event if (i) all descendants of the node ancestral to O (other than O) are Russian; (ii) one or more of immediate descendants of O are Russian singletons or Russian transmission lineages; and (iii) one of more of immediate descendants of O are clades consisting of only non-Russian sequences (Fig S2 d).

The above definitions are rather conservative and underestimate the true number of IBTs and OBTs. We additionally inferred migration events using Treetime (Figure S4). We labeled every sample on the tree as either Russian or non-Russian, and reconstructed the states of internal nodes for this trait. IBTs were then defined as edges whose parent and child nodes were inferred as non-Russian and Russian, respectively, with probabilities of at least 0.8. Groups of edges descendant to the same parent node were considered to constitute one IBT event. OBTs were defined analogously.

Data on monthly passenger traffic were obtained from the Federal Air Transport Agency webpage^9^.

### Validation of OBT events

Transmissions to New Zealand were cross-referenced with the local Christchurch cluster by sequence metadata (date and town of sample collection). No other cases were identified in Christchurch at that time. Information about transmissions to South Korea was extracted from GISAID metadata.

## Data Availability

All primary sequencing data obtained in this study together with corresponding metadata is being deposited to GISAID.

## Acknowledgements

This study was partially supported by RFBR project 20-04-60556. We thank Sergei L Kosakovsky Pond for help with HyPhy analyses, and Evgeniya Alekseeva and members of the Bazykin lab for fruitful discussions. We thank all of the authors who have contributed genome data on GISAID (see Supplementary Materials 2 for the list).

