## Supplementary for "Genomic epidemiology of SARS-CoV-2 in Russia reveals recurring cross-border transmission throughout 2020"

### Supplementary Figures

**Fig. S1. Prevalence of major PANGOLIN lineages[10] in Russia by period: A, March-July; B, August-November.**

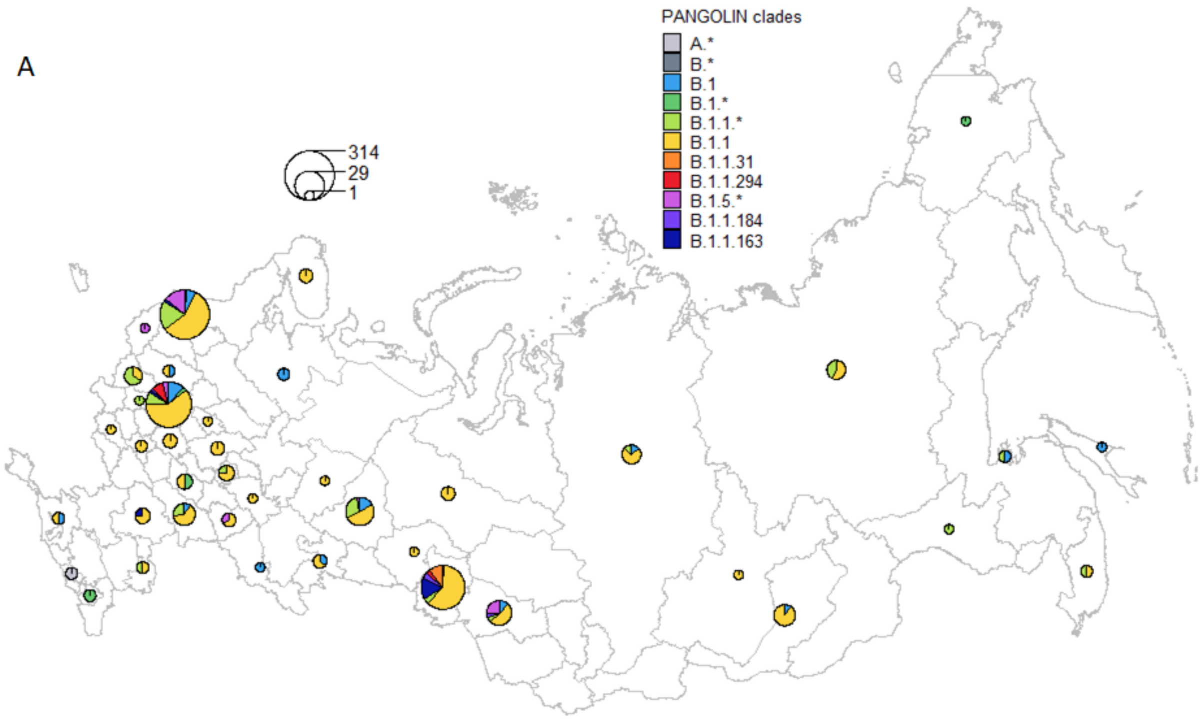

B

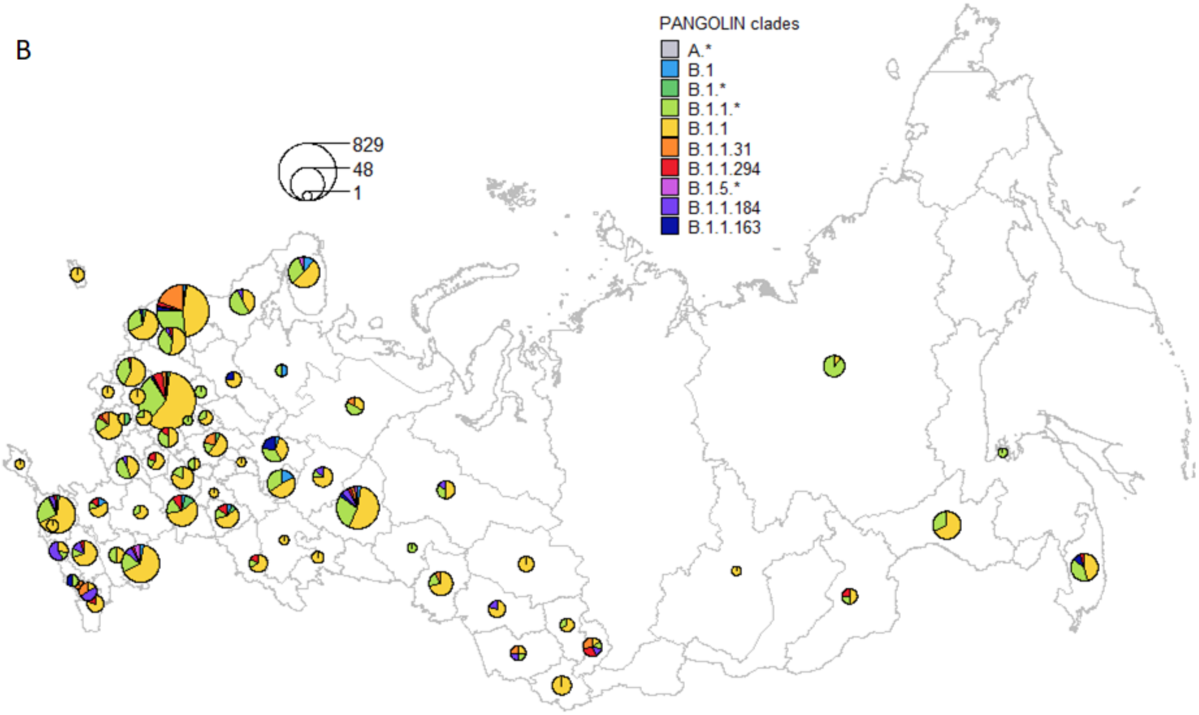
PANGOLIN clades
A.*
B.1
B.1.*
B.1.1.*
B.1.1
B.1.1.31
B.1.1.294
B.1.5.*
B.1.1.184
B.1.1.163
829
48
1

**Fig. S2. Phylogenetic categorization of sequences into Russian transmission lineages, stem clusters, stem-derived singletons (a) and singletons (b), and inference of IBTs (c) and OBTs (d).** I, node ancestral to an IBT event; O, node ancestral to an OBT event.

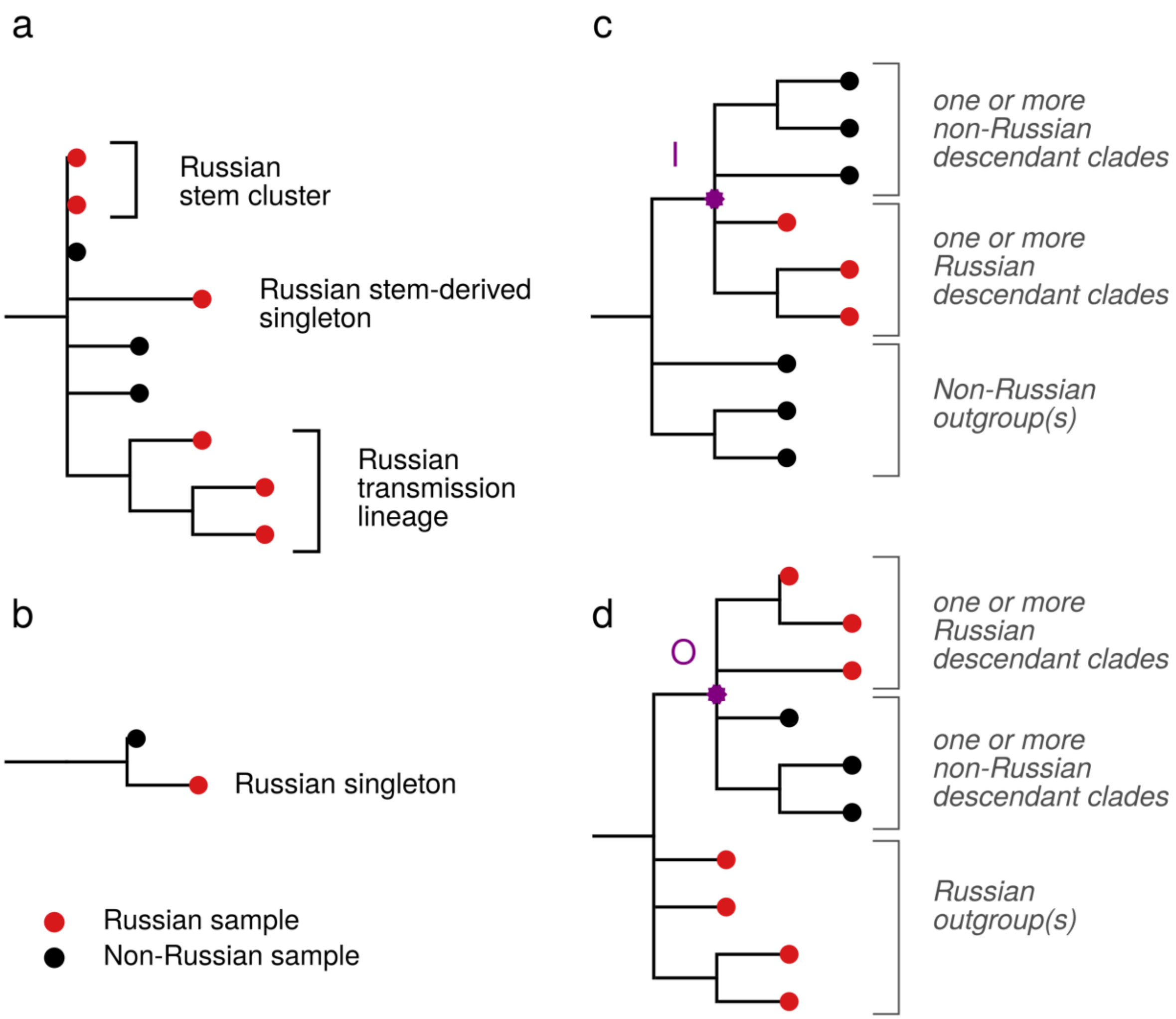

**Fig. S3. Rarefaction curves for numbers of inferred Russian transmission lineages.** The sequences from Moscow (A), Saint Petersburg (B) and all of Russia (C) were subsampled 10,000 times to the number shown on the horizontal axis, and the number of Russian transmission lineages was inferred. The shaded area shows the range of observed values.

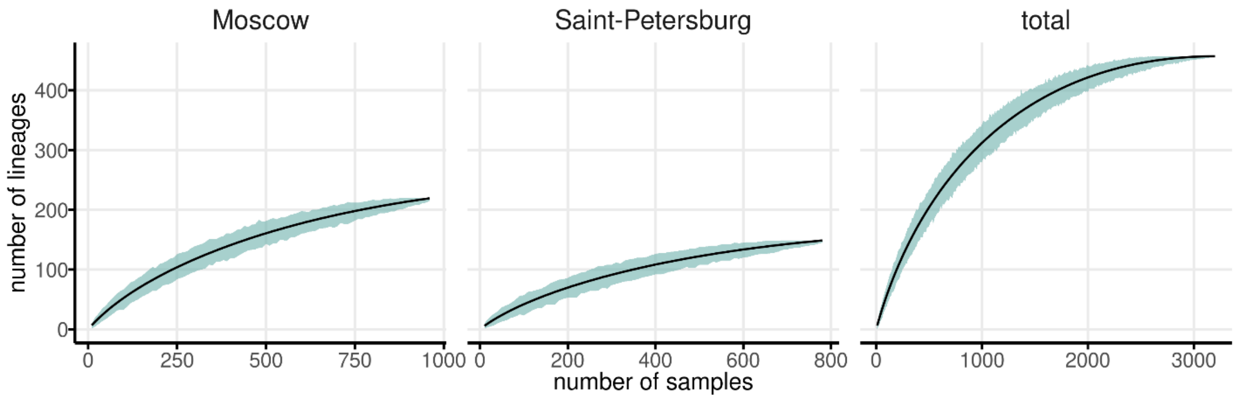

**Fig. S4. Timeline for inbound and outbound transmissions of SARS-CoV-2 variants, estimated by Treetime.** Notations are as in Fig. 8.

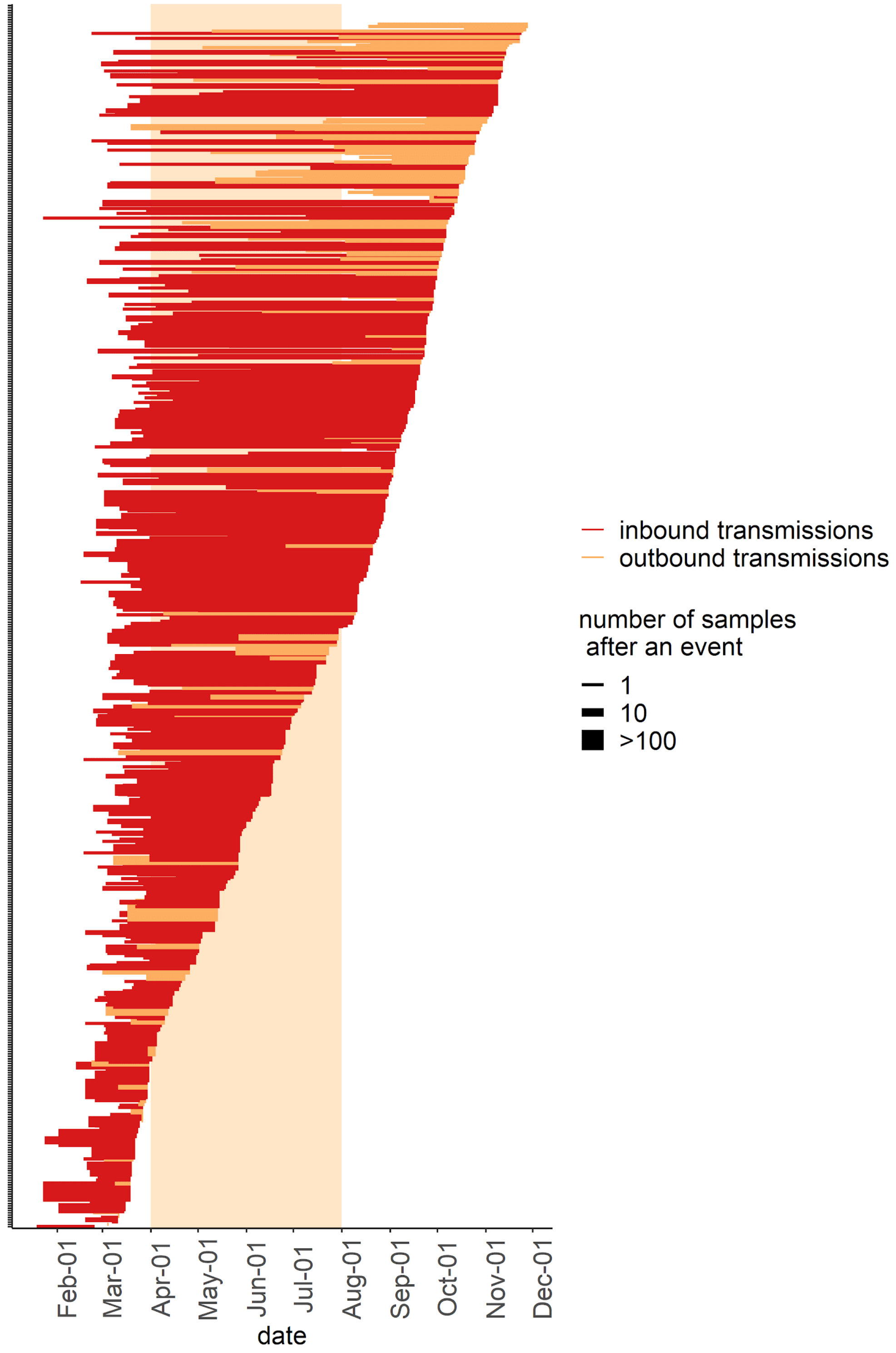

**Fig S5. OBTs to New Zealand.** Each tree represents an independent introduction event that occurred in the middle of October, each resulting in 6 sampled sequences. Branch lengths are measured in the number of changes. Samples from Russia are identified with red labels. OBTs are indicated by purple star and "O" letter.The number at the end of the sequence id represents the number of identical sequences (including the one shown) identified in the region on this date.

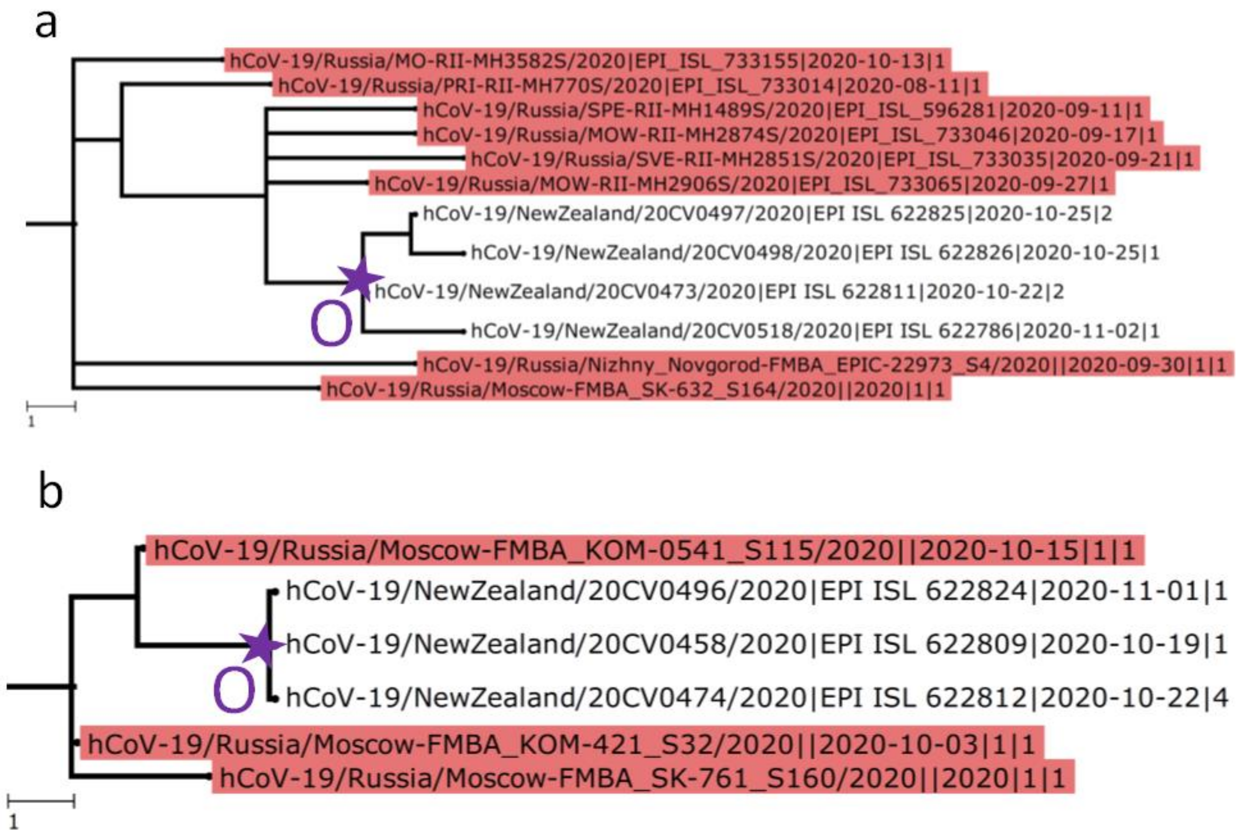

**Figure S6. OBTs to South Korea.** Notation as in Fig. S5.

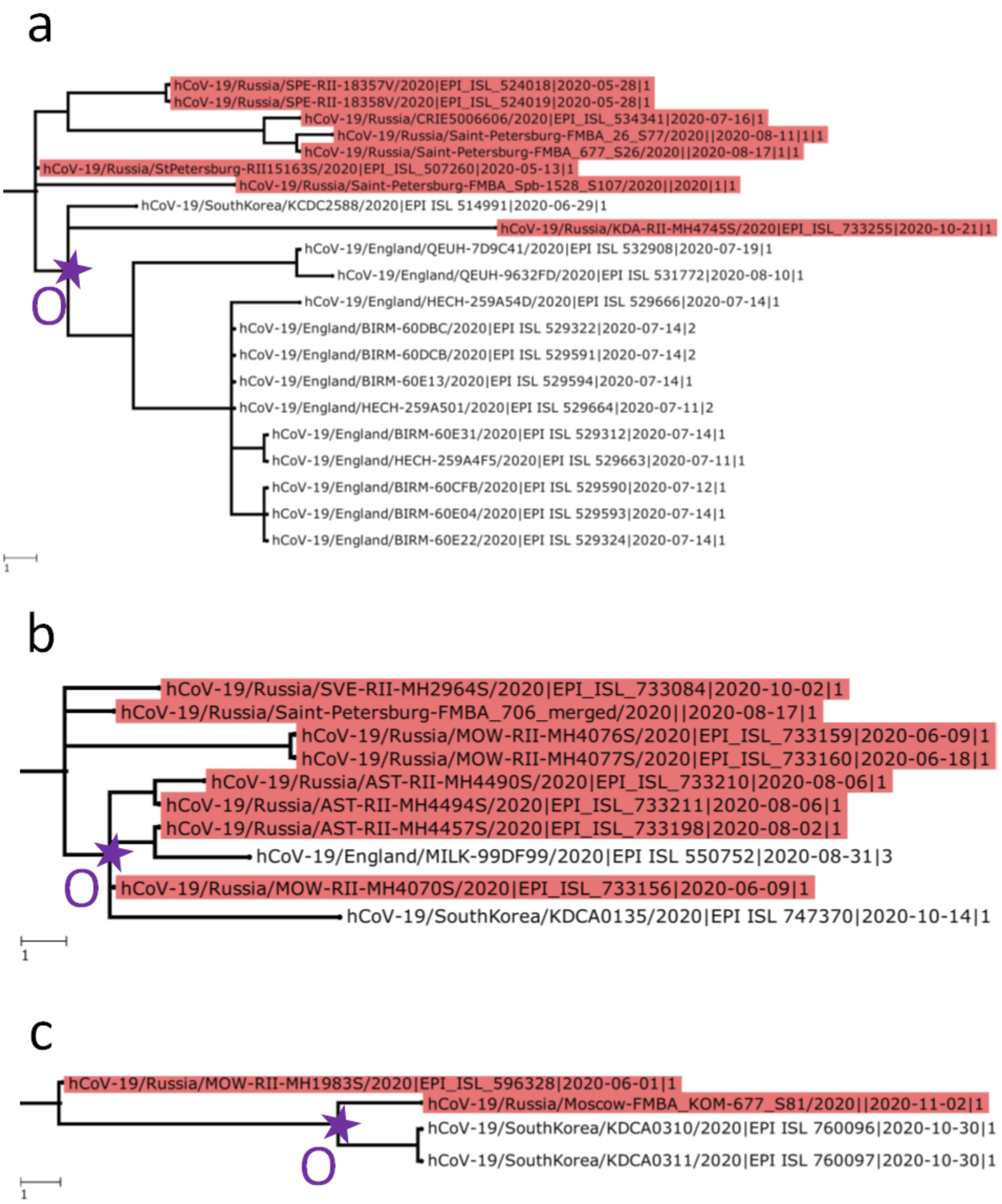

**Figure S7. Schematic representation of five distinct SARS-CoV-2 clades analysed independently in the work.** B.x consists of the B lineage and all its descendants except B.1 and lineages descendant from it; similarly, B.1.x includes B.1 and all its descendants but B.1.1 lineage and GH clade (GISAID nomenclature; denoted here as B.1.GH); B.1.1 and B.1.GH clades are analyzed separately. Sites carrying the key mutations defining the specified clades are indicated.

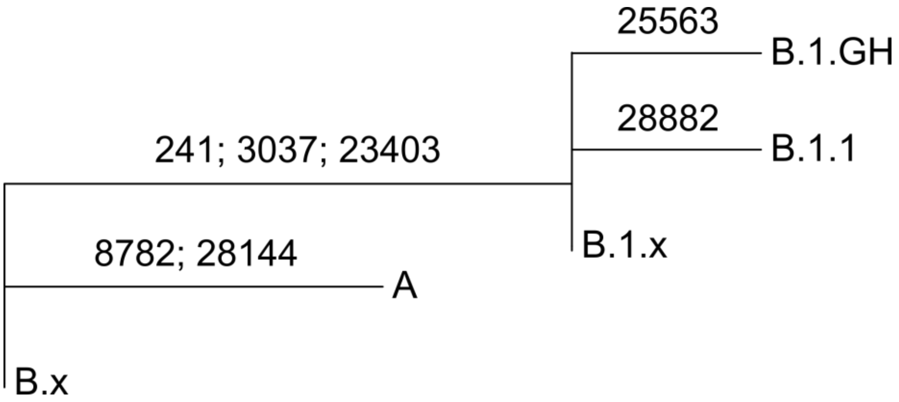
